# Salivary detection of COVID-19. Clinical performance of oral sponge sampling for SARS-CoV-2 testing

**DOI:** 10.1101/2021.02.17.21251556

**Authors:** Charles Hugo Marquette, Jacques Boutros, Jonathan Benzaquen, Marius Ilié, Mickelina Labaky, Didier Benchetrit, Thibaut Lavrut, Sylvie Leroy, Richard Chemla, Michel Carles, Virginie Tanga, Charlotte Maniel, Olivier Bordone, Maryline Allégra, Virginie Lespinet, Julien Fayada, Jennifer Griffonnet, Véronique Hofman, Paul Hofman

## Abstract

**Background:** The current diagnostic standard for coronavirus 2019 disease (COVID-19) is reverse transcriptase-polymerase chain reaction (RT-PCR) testing with naso-pharyngeal (NP) swabs. The invasiveness and need for trained personnel make the NP technique unsuited for repeated community-based mass screening. We developed a technique to collect saliva in a simple and easy way with the sponges that are usually used for tamponade of epistaxis. This study was carried out to validate the clinical performance of oral sponge (OS) sampling for SARS-CoV-2 testing.

**Methods:** Over a period of 22 weeks, we collected prospectively 409 paired NP and OS samples from consecutive subjects presenting to a public community-based free screening center. Subjects were referred by their attending physician because of recent COVID-19 symptoms (n=147) or by the contact tracing staff of the French public health insurance since they were considered as close contacts of a laboratory-confirmed COVID-19 case (n=262).

**Results:** In symptomatic subjects, RT-PCR SARS-CoV-2 testing with OS showed a 96.5% (95%CI: 89.6-94.8) concordance with NP testing, and, a 93.3% [95%CI: 89.1-97.3] sensitivity. In close contacts the NP-OS concordance (93.8% [95%CI: 90.9-96.7]) and OS sensitivity (71.9% [95%CI: 66.5-77.3]) were slightly lower.

**Conclusion:** These results strongly suggest that OS testing is a straightforward, low-cost and high-throughput sampling method that can be used for frequent RT-PCR testing of COVID-19 patients and mass screening of populations.

**Summary of the “take home” message:** OS sampling for SARS-CoV2 RT-PCR is an easy to perform, straightforward self-administered sampling technique, which has a sensitivity of up to 93.3% in symptomatic patients and 71% in close contact subjects.

## Introduction

To date, reverse transcriptase-polymerase chain reaction (RT-PCR) testing of naso-pharyngeal swab specimens (NP) is the gold standard for diagnosis of coronavirus 2019 disease (COVID-19) [1–4]. While its specificity is 100%, its sensitivity is 80-90% during the first 10 days of the disease of a COVID-19 positive subject [5, 6] and depends on the operator and on the moment the sampling is performed during the course of the infection. RT-PCR on NP, known as being of “high analytical sensitivity", is particularly well suited for symptomatic patients. However, in the context of a pandemic, when mass screening or repeated testing is required, the use of NP for RT-PCR is not appropriate [7] since its acceptability is debated and its implementation requires time and dedicated trained staff.

Antigen-detecting rapid diagnostic tests (Ag-RDT) are quick, simple, inexpensive and allow the decentralization of testing of symptomatic people at the point of care. As for a conventional RT-PCR test, they are performed with NP and thus need to be done by a trained operator [8]. The sensitivity varies considerably (60 to 95%) [8–10] and the invasiveness makes Ag-RDT poorly suited for repeated testing or mass screening.

Sampling of saliva offers a promising alternative to NP [11]. Various pathophysiological pathways explain the presence of the SARS-CoV-2 in saliva of patients with COVID-19 [12–17]. Tests with saliva involve amplification of viral RNA by RT-PCR or by Reverse Transcriptase Loop-Mediated Isothermal Amplification (RT-LAMP). These tests are non-invasive, easily repeatable and can be performed with or even without assistance, leading in the latter case to a lower risk of contamination of nursing staff. RT-PCR on saliva samples has been approved in Japan since June 2020 and in the USA since October 2020. Following the publication of an umbrella review [18] including ten meta-analyses [11, 19–27] as well as trials meeting predefined quality criteria, the French “Haute Autorité de Santé” [28] retained very recently the indication of RT-PCR on saliva in the following three situations: 1 / on symptomatic patients as a second-line alternative to NP when NP is difficult or impossible (deviation of the nasal septum, very young patients, patients with psychiatric disorders, etc.); 2 / on contact persons as a second-line alternative to NP for whom NP is difficult or impossible; 3 / in the first instance as part of targeted mass screening, particularly if it is repeated regularly such as in schools, universities, for staff in health care establishments, in nursing homes, etc … From a practical point of view, saliva sampling should be performed more than 30 minutes after the last drink, food, cigarette/e-cigarette, tooth brushing or mouth rinsing [28]. Saliva spit in a dry and sterile bottle is preferred; otherwise, saliva is collected using a dedicated system. Based on our research on the respiratory epithelium and on COVID-19 [29–33] we aimed to investigate the potential use of the RT-PCR detection of SARS-COV-2 on oral sponge samples (OS) as an alternative to NP for symptomatic subjects and close contacts.

## Methods

### Study design and participants

The present study was conducted on a prospective cohort of consecutive volunteers at the Nice-Côte d’Azur Metropolis community-based COVID-19 center (Nice, France), accessible for free screening to the general population.

During the first part of the study (Sept 21, 2020 to Jan 6, 2021) we enrolled subjects referred by their attending physician because of recent (≤ 2 weeks) symptoms of COVID-19 [34]. During the second part of the study (Feb 6 to March 8, 2021) we enrolled subjects referred by the contact tracing staff of the French public health insurance [35], since they were considered as close contacts of a laboratory-confirmed COVID-19 case (figure 1).

**Figure 1:**
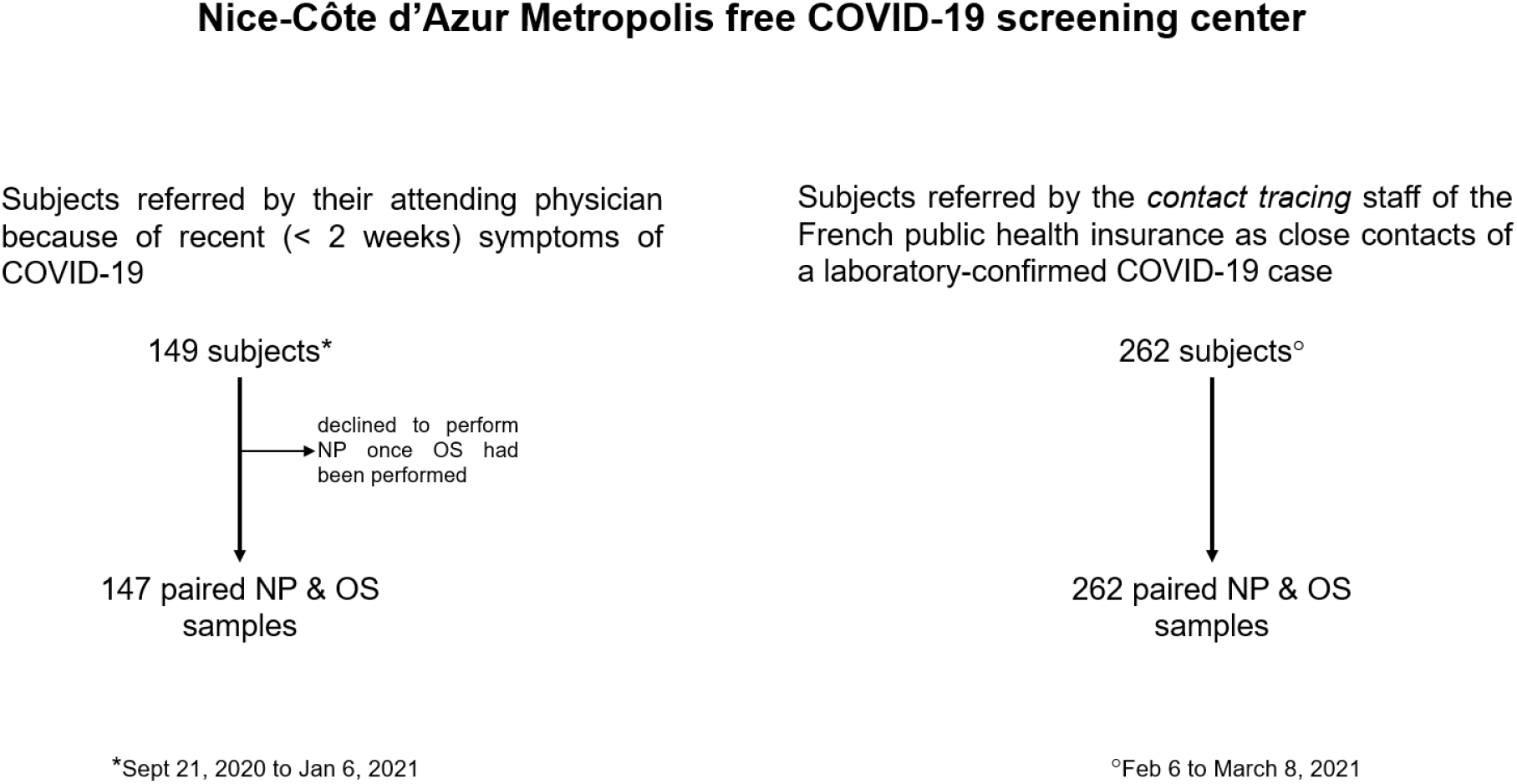
Trial profile

### Procedures

#### Specimen Collection

After signing an informed consent to participate, all volunteers were interviewed and underwent paired sampling: NP and OS in a random order. Disposable NP nylon swabs (type A-04. Jiangsu Han-Heng Medical technology Co., Ltd. Changzou, Jiangsu, China) were eluted into a vial containing viral transport medium (appendix 1) and transported within 4 hours to the Synlab Barla laboratory (Nice, France). Small hydroxylated polyvinyl acetate (PVA) sponges (Merocel® Standard Dressing, réf 400400, Medtronic), usually used for tamponade of epistaxis, were placed in the mouth by the subjects themselves and taken out after one minute and inserted into a sterile collection tube transported within 4 hours to the Idylla platform (Biocartis, Mechelen, Belgium) of the Laboratory of Clinical and Experimental Pathology (LPCE), (Hôpital Pasteur 1, Nice). No instructions were given regarding the need to fast, to have chewed gum or smoked a cigarette/e-cigarette.

#### Detection of SARS-CoV-2 RNA on NP samples

The Synlab Barla laboratory is a private biological analysis laboratory that is under contract with the Nice-Côte d’Azur Metropolis for COVID screening tests. This laboratory used RT-PCR to detect SARS-CoV-2 RNA on NP during the first part of the study. For logistical reasons, this laboratory switched to the Transcription Mediated Amplification (TMA) technique to detect SARS-CoV-2 RNA during the second part of the study. According to the guidelines of the French society for Microbiology [36] for RT-PCR, the result was considered positive when the Ct (cycle threshold) value for the N and/or on Orf1b genes was equal or less than 36. For TMA the result was considered as positive when the RLU was above 850.

#### Analysis of OS samples

We described previously RT-PCR for detection of SARS-CoV-2 RNA on the Idylla platform and showed a 100% positive and negative agreement between this technique and standard-of-care RT-PCR tests [33]. Upon arrival of the sample, 200μl of the OS eluate were pipetted with a sterile tip (D. Dutscher, reference 134000), placed individually into Idylla SARS-CoV-2 test cartridges (Biocartis, reference A1043/6) and underwent fully automated nucleic acid testing including extraction, amplification, and detection in a single-use cartridge. The duration of the run took 90 minutes (+/-10 min). The residual volume OS eluate was immediately aliquoted and stored at −80°C in the Nice COVID-19 Biobank, which is S96-900 certified by AFNOR [30]. According to the manufacturer, the Idylla SARS-CoV-2 test provides a qualitative result for the presence or absence of SARS-CoV-2 RNA with a corresponding quality status. The Biocartis SARS-CoV-2 test included 2 genes (N, Orf1b) covered by 5 PCR targets (2 N targets and 3 Orf1b targets). A positive result required at least 2 amplified N targets [by setting a quantification cycle (Cq) of 41.9] and/or at least one or more amplified Orf1b targets. As Orf1b is highly specific no threshold was required for this gene.

#### External validation

All OS eluates were sent to the Synlab Barla laboratory and underwent RT-PCR using the ORF1ab and N genes Da An Gene DA0992-Detection Kit for 2019-nCoV (Da An Gene Co., Ltd. Sun Yat-sen University, Guangzhou, Guangdong, China). During the first part of the study this external validation was performed on the OS eluates stored at −80°C in the Nice COVID-19 biobank) [30]. During the second part of the study, external validation was carried out in the Synlab Barla laboratory on fresh samples transferred from the Nice COVID-19 biobank.

The different analyses were processed in a double-blind way: the results of the reference standard test were unavailable to the readers of the index test and vice versa.

### Outcomes

The primary outcome was the clinical performance of OS compared to NP for SARS- CoV-2 screening in symptomatic and close contact subjects. For each group (symptomatic vs close contacts) we extracted the number of individuals positive for SARS- CoV-2 in both NP and OS (a), those positive only with NP (b), those positive only with OS (c) and (d) those negative with both NO and OS (Table 1). From these data, we calculated the concordance of the two types of sample (a+d)/(a+b+c+d). We also calculated the sensitivity of the test on each type of sample. The estimation of the sensitivity of a test requires a reference diagnosis. Since NP sampling has been shown to produce false negatives by RT-PCR, sensitivities for NP and OS are defined here respectively as (a+b)/(a+b+c) and (a+c)/(a+b+c), considering as a true positive any individual with a positive result on one or the other sample. This definition of a positive individual is in agreement with the US-CDC and the ECDC directives on SARS-CoV-2 testing [37, 38].

**Table 1a:** Clinical performance of oral sponge (OS) compared to nasopharyngeal swab (NP) for SARS- CoV-2 screening. LPCE, Idylla platform.

|  | NP +<br>OS + | NP +<br>OS - | NP -<br>OS + | NP -<br>OS - | Total | Concordance | Sensitivity of NP | Sensitivity of OS |
| --- | --- | --- | --- | --- | --- | --- | --- | --- |
|  | a | b | c | d | n =<br>a+b+c+d | (a+d)/n<br>[95% CI] | (a+b)/(a+b+c)<br>[95% CI] | (a+c)/(a+b+c)<br>[95% CI] |
| Symptomatic subjects | 54 | 4 | 1 | 88 | 147 | 96.5%<br>[93.5 – 99.5] | 98.3%<br>[96.2 - 100] | 93.2%*<br>[89.1 - 97.3] |
| Close contact subjects | 41 | 16 | 0 | 205 | 262 | 93.8%<br>[90.9 – 96.7] | 100% | 71.9%<br>[66.5 – 77.3] |

**Table 1b:**
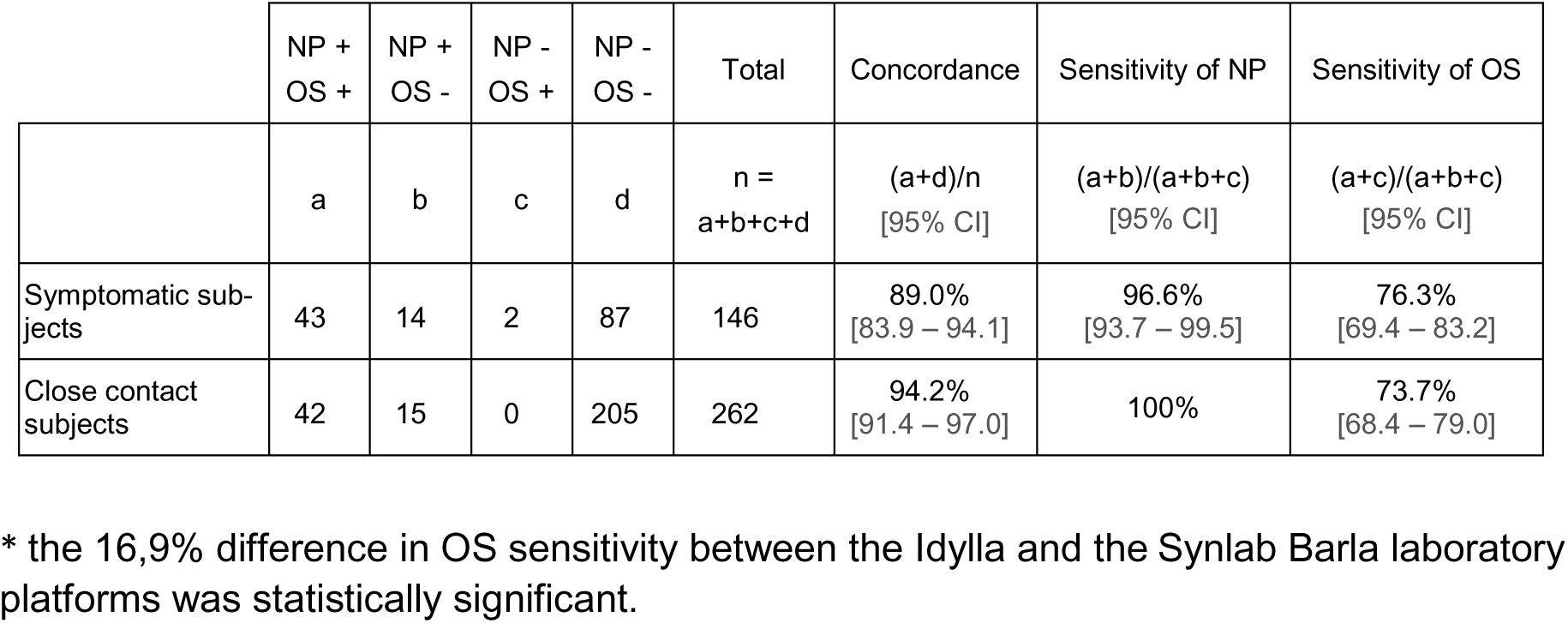
Clinical performance of oral sponge (OS) compared to nasopharyngeal swab (NP) for SARS- CoV-2 screening. External validation at the Synlab Barla laboratory platform

Secondary outcomes included the description of the studied population and the comparison between Ct values measured with NP and OS, an approximate proxy of viral loads. To calculate the sample size in the absence of data regarding the sensitivity of OS in symptomatic subjects, we calculated the sensitivity of OS once the first 40 paired (NP/OS) samples were obtained. This sensitivity was 85%. Given this estimate and a desired lower bound of the 95% confidence interval for sensitivity of at least 80%, 144 volunteers were needed to complete the study in symptomatic subjects. To deal with the dropout risk we decided to include 149 volunteers.

### Statistics

Continuous variables are presented as means (± SD), and categorical variables as numbers and percentages. Baseline characteristics between patients with and without COVID-19 were compared using the Student’s t-test or Wilcoxon - Mann Whitney for quantitative variables based on the normality of the distribution of parameters or using the Chi-Square test for qualitative variables.

### Ethics and regulatory authorizations

The promoter of the study was the Center Hospitalier Universitaire de Nice. The agreement for the study of the Institutional review board Sud Méditerranée V was obtained on April 22, 2020 (registration # 20.04014.35208). SHAM liability insurance (n° 159087). The study is registered in ClinicalTrial.gov (NCT04418206).

### Role of the funding sources

The organizations that supported this study played no role in its design, patient selection, data collection, analysis or interpretation, report writing, or decision to submit the document for publication. Authors had full access to all data and responsibility of submission for publication.

## Results

Four hundred and eleven subjects underwent COVID-19 diagnostic testing over the 22 weeks of the study, 149 during the first part of the study conducted on symptomatic subjects and 262 during the second part of the study conducted on close contacts. Two symptomatic subjects declined to perform NP once OS had been performed and were excluded from subsequent analyses. One had a positive OS RT-PCR, and the other was negative. The study therefore covers the remaining 409 participants (figure 1).

The 147 symptomatic subjects were predominantly women 86/147 (58.5%). Their mean age was 40 ± 15 years. The interval between symptom onset and testing was 3.6 ± 2.6 days and most participants (107/147 [72.8%]) were sampled at the early stage of the disease, i.e., within 4 days of symptom onset. Of these symptomatic subjects, 59 (40.1%) had a positive RT-PCR result with one or both of the sampling techniques and were thus diagnosed COVID-19. No clinical symptoms distinguished between RT-PCR positive and negative subjects (figure 2), with the exception of anosmia and dysgeusia, which were more frequent in RT-PCR positive subjects (42% vs. 10%, p <0.001 and 38 vs. 16%, p = 0.004) and sore throat, which was significantly more common in RT-PCR negative subjects (36 vs. 10%, p = 0.001).

**Figure 2:**
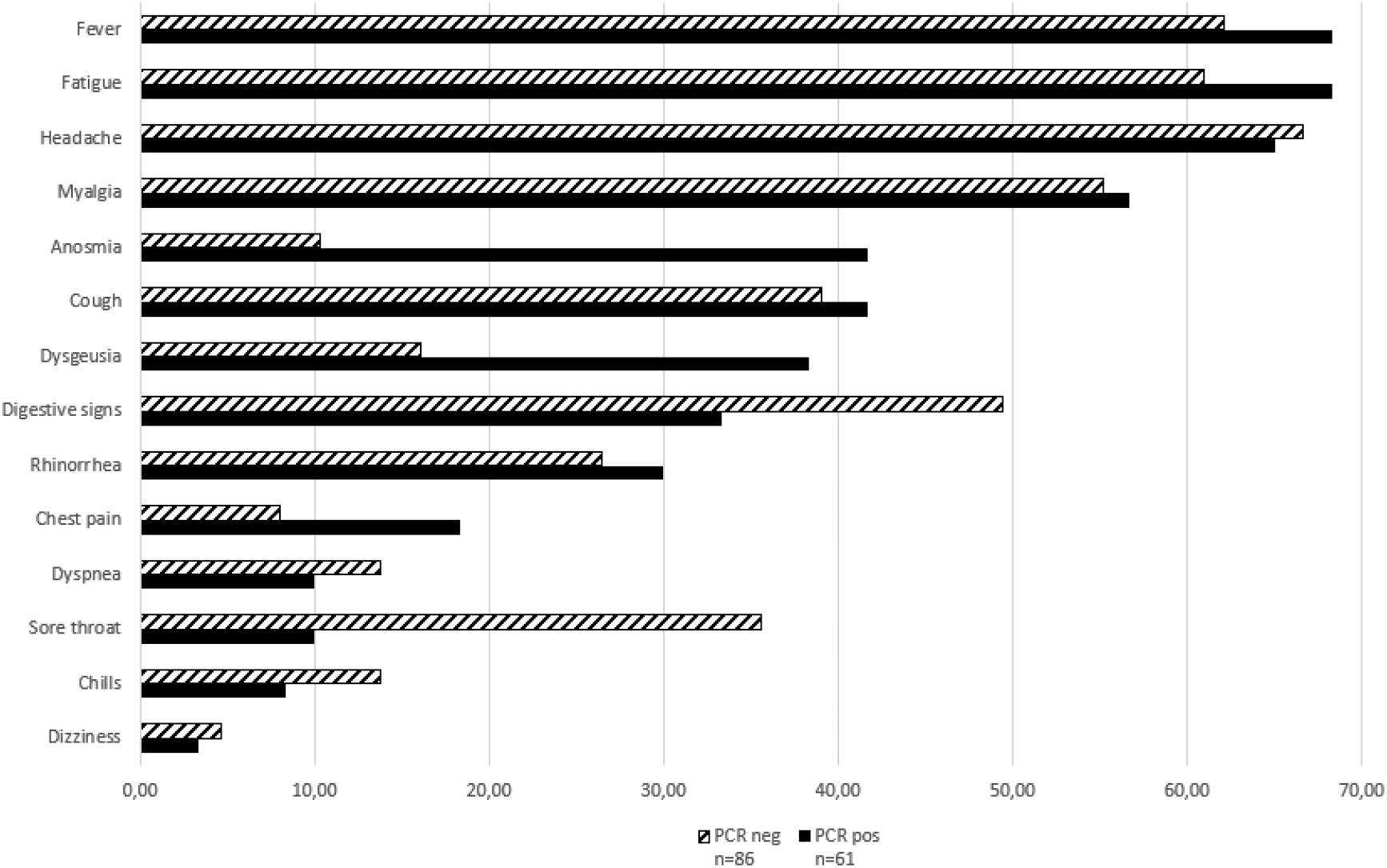
Self-reported symptoms (%)

**Figure 3:**
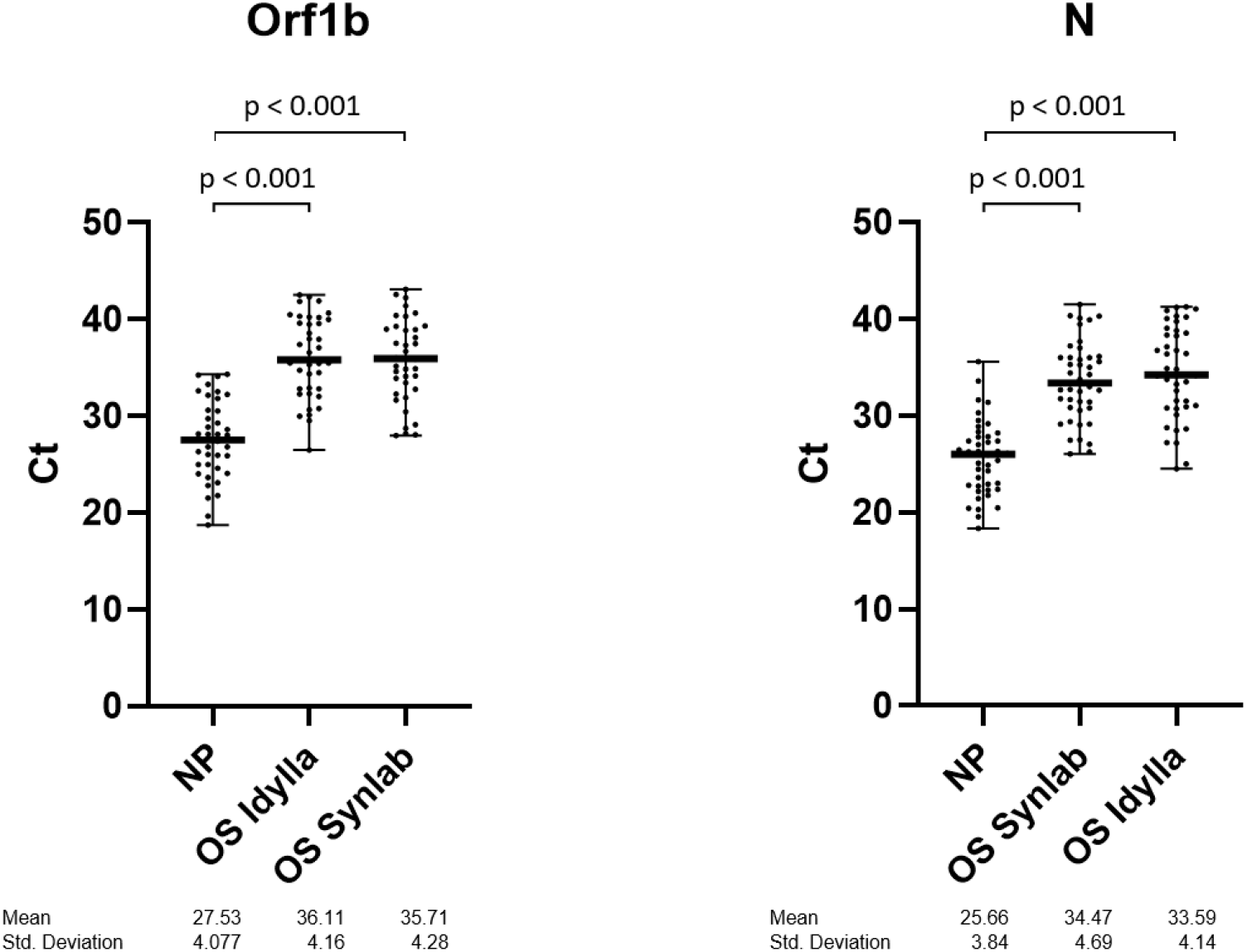
Viral load of NP and OS as indirectly assessed by the Ct for the Orf1b and N genes

For these symptomatic subjects, the Ct values for the Orf 1b and N genes with OS showed a more than 3-point increase compared with NP; the magnitude of this difference indicates a more than 10-fold lower quantity of viral genetic material in OS samples [39].

The 262 close contacts with a laboratory-confirmed COVID-19 case were predominantly women 145/262 (55.3%). The mean age was 41 ± 15 years. The contact was identified in the household (n=41; 15.6%), in the family (n=56; 21.4%); in the close environment (n=40; 15.3%), at work (n=65; 24.8%), or was not known by the subject (n=60; 22.9%). Of these close contact subjects, 57 (21.7%) had a positive RT-PCR result with one or both of the sampling techniques. Differences in viral load between the NP and the OS could not be assessed in close contact subjects since viral RNA detection with NP relied on TMA and not on RT-PCR in this part of the study.

In the symptomatic subjects, the NP-OS concordance was 96.5% (95%CI: 93.5 – 99.5) and the OS sensitivity was 93.2% (95%CI: 89.1 – 97.3) with the Idylla platform. With the Synlab Barla platform, the NP-OS concordance was 89.0% (95%CI: 83.9 – 94.1) and the OS sensitivity was 76.3% (95%CI: 69.4 – 83.2) (table 1).

In the close contact subjects, the NP-OS concordance was 93.8% (95%CI: 90.9 – 96.7) and the OS sensitivity was 71.9% (95%CI: 66.5 – 77.3) with the Idylla platform. With the Synlab barla platform, the NP-OS concordance was 94.2% (95%CI: 91.4 – 97.0) and the OS sensitivity was 73.7% (95%CI: 68.4 – 79.0) (table 1).

## Discussion

In this study, RT-PCR SARS-CoV-2 testing with OS showed a high level of concordance (with NP testing), 96.5% (95%CI: 89.6-94.8) and sensitivity, 93.3 (95%CI: 89.1-97.3) for symptomatic subjects and a slightly lower level of concordance, 93.8% (95%CI: 90.9-96.7) and sensitivity, 71.9% (95%CI: 66.5-77.3) in close contacts of a laboratory-confirmed COVID-19 case. These results obtained in ambulatory subjects presenting to a public community-based screening center, strongly suggest that OS, a straightforward, low-cost and high-throughput sampling material can be used for frequent RT-PCR testing of COVID-19 patients and mass screening of populations.

During the early phase of the COVID-19 pandemic, many molecular tests, immunoassays and sampling methods were rapidly developed and validated using archived biological samples of known virological status, albeit many still await clinical validation [40]. To evaluate the clinical performance of OS, compared to NP, the present study was carried out according to the standards recommended by the French “Haute Autorité de Santé” [8]; i.e., a prospective comparative clinical study relating to a series of individuals of unknown COVID-19 status, recruited consecutively; a salivary test including at least two molecular targets; and by the US and European CDC directives; i.e. definition of a positive individual when comparing two diagnostic SARS-Cov-2 testing methods [37,38].

In published studies that compared salivary and NP testing, symptomatic subjects were the dominant inclusion profile (85%), especially in the majority of short series of hospitalized patients [18]. Only 6 cohorts of equal or larger size than ours reported comparative test results obtained in a community-based settings [41–46].

Given the particularly high incidence of COVID-19 in the Nice metropolitan area throughout our study period (incidence between 330 and 720/100,000), our rate of positivity (21-41%) was higher than the rates observed in these 6 cohorts (1.5% to 10.7%). Despite this difference, our NP-OS concordance and the sensitivity of RT-PCR on NP and on OS are in agreement with the results reported for these cohorts; and with the notion of no association between salivary and NP test performance and viral prevalence [19].

RT-PCR with OS performed at the Synlab Barla platform showed a significantly lower sensitivity than Idylla platform in symptomatic subjects. This discrepancy may be due to 2 factors: the Idylla platform uses 3 Orf1 and 2 N targets while the Synlab Barla platform uses only one Orf1 and one N target; preservation at −80°C for several weeks may have altered the nucleic acids. The discrepancy disappeared in the second part of the study in which the external validation of OS testing at the Synlab Barla platform was done on fresh samples in real time.

The significant difference in viral load observed between samples of nasopharyngeal and buccal origin may be due to several parameters, notably endogenous to the laboratory [(i.e. total volume of sample collection buffer/medium, sample preparation method (heat, lysis methods)] and laboratory reagent volumes used in each step of the RT-PCR process. A similar difference has been observed by others [2, 47–49] and explains why the diagnostic yield of NP is considered to be higher than that of throat swabs [48, 49]. Thus, this difference most likely has a pathophysiological basis, related to the fact that the mouth squamous cells act as a viral reservoir to a lesser degree than the nasopharynx ciliated cells [31, 32, 50]. As a consequence, the interpretative cut-offs for NP Ct [36] hardly apply to saliva samples.

The OS sampling method is well standardized. It does not need sialagogic drugs, nor clearing the throat or spitting effort, nor particular constraints such as early morning saliva sampling before tooth brushing and breakfast, avoiding eating, drinking, gum chewing, smoking, or vaping. [51–53]. It does not need dedicated trained nursing staff and its acceptability makes it possible to consider repeating the test even in institutionalized elderly people or in children.

The question as to whether the 72% sensitivity we observed in contact subjects is good enough to identify SARS-CoV-2 infected people in the setting of mass screening has been debated by Mina et al. in a landmark position paper [7]. In this paper they distinguished between a low frequency tests with a high analytical sensitivity and high frequency tests with low analytical sensitivity. RT-PCR with NP samples belongs to the former category and is well suited to diagnosis of COVID-19 in symptomatic subjects. Whereas the OS sampling method described herein belongs to the latter category and would be more effective in identifying, isolating, and thus filtering out currently infected persons, including those who are asymptomatic.

We industrialized the OS process in the form of an “all-in-one” kit simplifying extraction of the biological liquid (figure 4) and presently intend to clinically validate the performance of this sampling method with the pooling technique [54] for mass screening.

**Figure 4:**
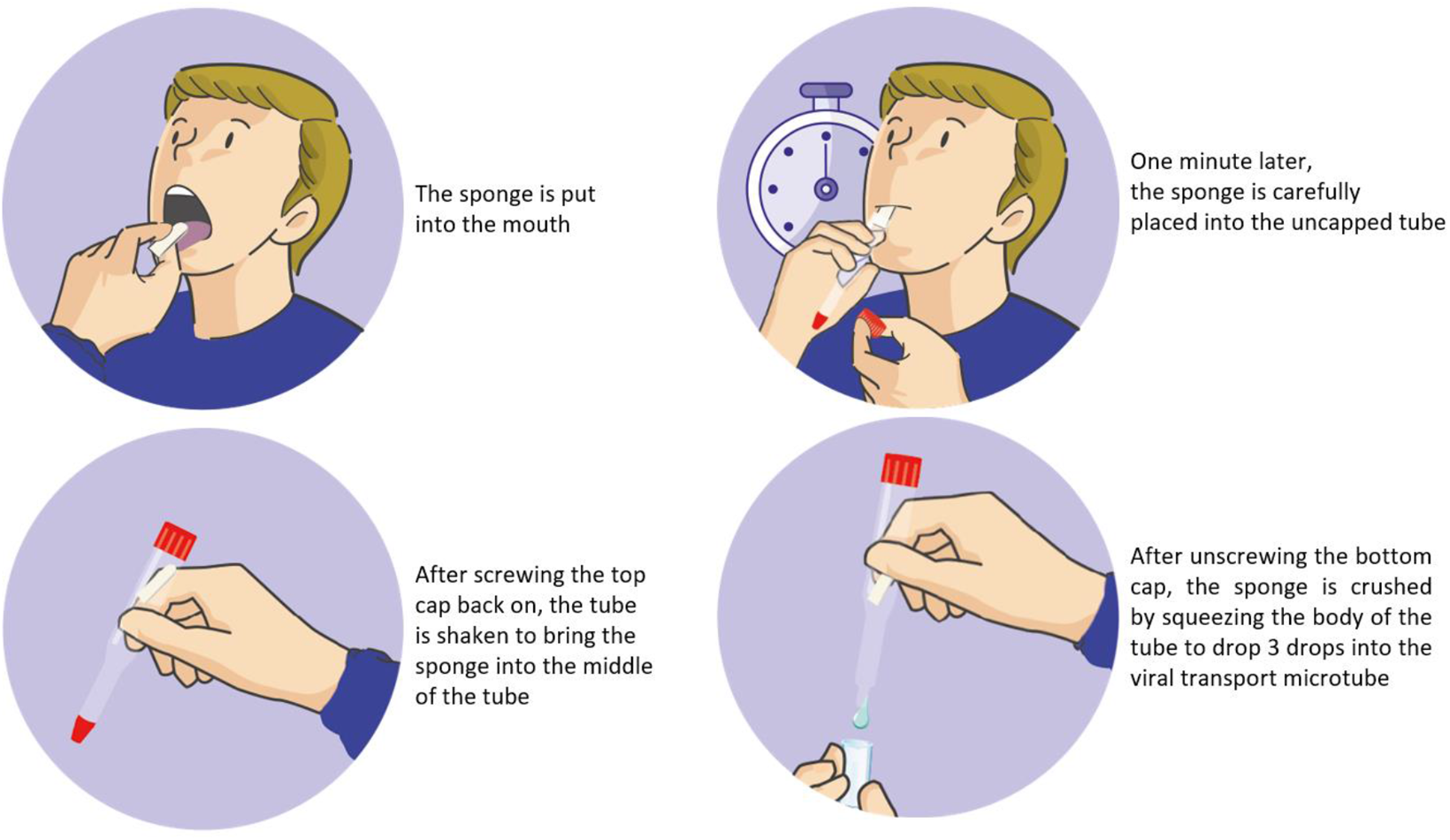
All-in-on saliva sampling technique with a PVA sponge and a flexible, double-capped plastic tube.

## Data Availability

all data referred to in this manuscript can be available after contacting the corresponding author

## Funding

This work was supported by Conseil Départemental 06, Ville de Nice, Métropole Nice Côte d’Azur, Fonds de Dotation AVENI, and from private donators.

## Acknowledgements

Special thanks to E. Faidhi, N. Fridlyand, A. Rauscher, E. Maris, the Lauro family and to the many private donators for their generous contribution.

## 1 APPENDIX PROCEDURES

### Detection of SARS-CoV-2 RNA with naso-pharyngeal swab specimens

During the first study period (September 21, 2020 to January 6, 2021; symptomatic patients), naso-pharyngeal (NP) swab specimens were analyzed by RT-PCR using the ORF1ab and N genes Da An Gene DA0992-Detection Kit for 2019-nCoV (Da An Gene Co., Ltd. Sun Yat-sen University, Guangzhou, Guangdong, China). NP swabs were eluted into a vial containing 400 µl of viral RNA extraction buffer (RNA/DNA purification kit, Da An Gene, ref DA0940); and 5 μl was then processed on an AGS 4800 Thermocycler (Hangzhou, Zhejiang, China). According to the guidelines of the French Society for Microbiology [reference 30 Avis du 25 septembre 2020 de la Société Française de Microbiologie (SFM) relatif à l’interprétation de la valeur de Ct] the result was considered as positive when the Ct (cycle threshold) value for the N and/or Orf1b genes was equal or less than 36.

During the second study period (February 6 to March 6, 2021; close contacts) NP samples were analyses with the Aptima^®^ SARS-CoV-2 assay (Hologic, Inc. 10210 Genetic Center Drive San Diego, CA 92121 USA), which combines Transcription Mediated Amplification (TMA), and Dual Kinetic Assay (DKA) that amplifies and detects two conserved regions of the ORF1ab gene. NP swabs were eluted into a vial containing 710 µl of medium (Hologic Specimen Lysis Tubes, ref PDD-06554) before being tested on the Panther system. Assay results were determined with a cut-off based on the total Relative Light Units (RLU). According to the guidelines of the French Society for Microbiology the results were considered as positive when the RLU was above 850.

### *Detection of SARS-CoV-2 RNA with oral sponge (OS) specimens at* the Synlab Barla laboratory

25 μl of the OS eluates transferred fresh from the Nice COVID-19 biobank were put into a vial containing 400 µl of viral RNA extraction buffer (RNA/DNA purification kit, Da An Gene, ref DA0940); and 5 μl was then processed on an AGS 4800 Thermocycler (Hangzhou, Zhejiang, China).

## Notes

### Competing Interest Statement

Paul Hofman is a member of the scientific advisory board (group of european experts) of Biocartis.  However, this boad is totally independ of Biocartis

### Clinical Trial

NCT04418206

### Funding Statement

This work was supported by Conseil Departemental 06, Ville de Nice, Metropole Nice Cote d'Azur, Fonds de Dotation AVENI, and from private donators.
Acknowledgements:
Special thanks to E. Faidhi, N. Fridlyand, A. Rauscher, E. Maris, the Lauro family and to the many private donators for their generous contribution.

### Author Declarations

The promoter of the study was the Center Hospitalier Universitaire de Nice. The agreement for the study of the Institutional review board Sud Mediterranee V was obtained on April 22, 2020 (registration # 20.04014.35208). SHAM liability insurance (n 159087).

### Summary of Updates

the legend in Table 1 contained errors that have been corrected in this version

